# Is there an airborne component to the transmission of COVID-19? : a quantitative analysis study

**DOI:** 10.1101/2020.05.22.20109991

**Authors:** Clive B. Beggs

## Abstract

**Objectives:** While COVID-19 is known to be spread by respiratory droplets (which travel <2m horizontally), much less is known about its transmission via aerosols, which can become airborne and widely distributed throughout room spaces. In order to quantify the risk posed by COVID-19 infectors exhaling respiratory aerosols in enclosed spaces, we undertook a computer modelling study to simulate transmission in an office building.

**Methods:** Respiratory droplet data from four published datasets were analysed to quantify the number and volume of droplets <100μm diameter produced by a typical cough and speaking event (i.e. counting from 1 to 100). This was used in a stochastic model to simulate (10000 simulations) the number of respiratory particles, originating from a COVID-19 infector, that would be inhaled in one hour by a susceptible individual practicing socially distancing in a 4 × 4 × 2.5m office space. Several scenarios were simulated that mimicked the presence of both symptomatic and asymptomatic COVID-19 infectors.

**Results:** On average, each cough and speaking event produced similar numbers of droplets <100μm diameter (median range = 971.9 – 1013.4). Computer simulations (ventilation rate=2AC/h) revealed that sharing the office space with a symptomatic COVID-19 infector (4 coughs and 10 speaking events per hour) for one hour resulted in the inhalation of 16.9 (25-75^th^ range = 8.1-33.9) aerosolised respiratory droplets, equating to about 280-1190 particles inhaled over a 35-hour working week. Sharing with an asymptomatic infector (10 speaking events per hour) resulted in the about 196–875 particles inhaled over 35 hours.

**Conclusions:** Given that live SARS-CoV-2 virions are known to be shed in high concentrations from the nasal cavity of both symptomatic and asymptomatic COVID-19 patients, the results suggest that those sharing enclosed spaces with infectors for long periods may be at risk of contracting COVID-19 by the aerosol route, even when practicing social distancing.

## 1.0 Introduction

Although it is widely acknowledged that COVID-19 is primarily transmitted by respiratory droplets [1], there is much less agreement as to whether or not the disease can also be transmitted by the airborne route [2]. Currently, the World Health Organization (WHO), while acknowledging that infectious respiratory droplets are widely liberated when COVID-19 patients cough, does not believe that airborne transmission plays any significant role in the transmission of COVID-19 [1]. Given that respiratory droplets travel through the air and are therefore technically ‘airborne’, this might appear at first sight to be a contradictory position to take. However, for many years the medical profession has drawn a clear distinction between diseases where transmission occurs via respiratory droplets impacting on the nasal and oral mucosae or conjunctiva, and those caused by the inhalation of small aerosol particles <5 or 10 μm in diameter (known as droplet nuclei) which are capable of reaching the alveoli in the lungs. The former involve large droplets, which once emitted are generally thought to fall rapidly to the floor, travelling <2 m horizontally from the source, whereas the latter, which includes tuberculosis (TB), is associated with droplet nuclei that are so small that they can remain suspended in air for hours, often travelling considerable distances. This distinction is important, because it forms the basis of the WHO social distancing guidelines regarding COVID-19, that require the general public should maintain a distance >1 m between individuals [1].

While the above distinction between droplet transmission and airborne transmission is widely used in medical texts, it is primarily based on clinical disease characteristics rather than on any principles of physics or aerosol science. As such, there are many inconsistencies in the position held by the WHO, and this has caused several research teams to challenge the assumption that airborne transmission does not contribute towards the spread of COVID-19 [3-8]. Chief amongst these inconsistencies is the false assumption that a clear-cut boundary exists between droplets and droplet nuclei [8-10]. In reality there is no clear demarcation between respiratory droplets and droplet nuclei - rather, there is a continuum, with the droplets rapidly reducing in size due to evaporation, with many becoming droplet nuclei that can remain suspended in the air for considerable periods of time [6, 11]. For example, it has been shown that droplets as large as 100 μm will evaporate in normal room conditions to become droplet nuclei before they can reach the floor [12-14]. Consequently, relatively large respiratory droplets, which may contain many viral particles, can rapidly reduce in size to become small droplet nuclei, which behave as aerosol particles and thus can be widely dispersed on room air currents. Indeed, some researchers investigating airborne transmission of respiratory viruses have found the greatest viral load to be associated with small droplet nuclei. For example, Bischoff et al [15] found that up to 89% of influenza virus-carrying particles in a healthcare setting were <4.7 μm in diameter, while Lindsley et al [16], investigating cough droplets produced by influenza patients, found 65% of the viral RNA to be in particles <4 μm, with airborne RNA recovered from 81% of the patients tested. Others have found similar results [17, 18]. Viral RNA was also recovered from aerosols during the 2003 epidemic of severe acute respiratory syndrome (SARS) [19]. Given that SARS is closely related to COVID-19, and the fact that approximately 99% of the particles produced during coughing end up as droplet nuclei <10 μm diameter [20], there is need to better understand the role that aerosol particles might play in the transmission of COVID-19.

While numerous texts refer to large droplets and small droplet nuclei, there is considerable confusion in the literature as to what actually constitutes each of these two classes. Earlier researchers, Wells [12] and Duguid [21] defined large droplets as having a diameter >100 μm, very much in line with the experimental work relating to droplet evaporation [13]. However, in more recent years >10 μm has often been used [10, 22, 23], although >5 μm has now become the accepted criteria [1, 24]. This classification being primarily based on the fact that droplet nuclei must be <10 μm in order to reach the alveoli in the lungs [10]. As such, this has led to the ludicrous situation where respiratory particles of say 15 μm diameter, which take approximately 5 minutes to fall 2 m is still air and thus can be easily transported on room air currents [25, 26], are classified as large droplets and wrongly assumed not to become airborne. As a result, many incorrect ideas have crept into the literature that have influenced the scientific debate on COVID-19. For example, the WHO state:

*“According to current evidence, COVID-19 virus is primarily transmitted between people through respiratory droplets and contact routes* … *Airborne transmission is different from droplet transmission as it refers to the presence of microbes within droplet nuclei, which are generally considered to be particles <5 μm in diameter, can remain in the air for long periods of time and be transmitted to others over distances greater than 1 m.”* [1]

Even more starkly, Gralton et al [24] state:

*“Droplet transmission is associated with particles sized >5 μm in diameter and airborne transmission is associated with particles sized ≤5 μm in diameter. ”* [24]

Here the WHO classify all respiratory droplets >5 μm as being ‘large’, implying that they cannot travel any great distance in the air. However, while this might be true for droplets >100 μm diameter, it is certainly not the case for all droplets >5 μm [13, 25, 26], many of which can remain suspended in the air for considerable periods of time and may contain SARS-CoV-2 viral particles [6]. Consequently, there is much confusion in the literature, as highlighted in a recent report of a COVID-19 outbreak in a crowded in a restaurant in Guangzhou, China [27], where, at the same time as ruling out the possibility of airborne transmission, it was reported that the outbreak was caused by droplet transmission ‘prompted’ by air-conditioned ventilation. This ambiguous conclusion, raises intriguing questions: were the air conditioning ‘jets’ strong enough to transport very large respiratory droplets >100 μm diameter, or was it the case that the droplets were say only 20 – 40 μm and thus relatively easy to transport? Given that rapid evaporation of the droplets would almost certainly have occurred [13, 25], it is highly likely that the ‘droplets’ that infected individuals in this outbreak were much less than 100 μm in diameter, in which case they could have become truly airborne as implied in subsequent analysis of the outbreak [3]. As such, it may be the case that airborne transmission of COVID-19 is being ruled out due to an incorrect understanding of the definition of ‘airborne’, and that in fact the aerial dissemination of respiratory droplets <100 μm in diameter is contributing to the transmission of COVID-19 in enclosed spaces. Indeed, recent work demonstrating that coughs and sneezes produce gas plumes [28] that can transport droplets and droplet nuclei many meters [5, 11], suggests that droplet nuclei containing SARS-CoV-2 virions might be widely distributed in enclosed spaces if infectors are present.

Giving the considerable ambiguity surrounding the transmission of COVID-19 by aerosol particles, we designed the computer modelling study reported here in order to estimate the contribution that airborne transmission might play in the spread of COVID-19 in buildings. The study was not intended to be definitive; rather, given the many unknowns that exist, it was designed simply to yield a first approximation that might be useful to epidemiologists and public health authorities when making decisions regarding the control of COVID-19.

## 2.0 Methods

Given that it is an emerging infectious disease that has only recently been identified, no data exist regarding the size distribution of droplets produced by symptomatic and asymptomatic COVID-19 patients. We therefore decided to use existing published data on respiratory exhalation events (REEs) in our models; that is, data relating to the liberation of respiratory droplets and droplet nuclei during a single cough and when speaking (i.e. counting from 1 to 100). To this end, we used datasets acquired by Loudon and Roberts [29], Duguid [21], Chao et al. [30], and Xie et al. [31] relating to droplet size distributions produced by healthy individuals when speaking and during a single cough. These four datasets were utilized because they all measured the droplet production associated with speaking by asking subjects to count from 1 to 100, and as such, were broadly comparable. Furthermore, with exception of Loudon and Roberts [29], all the researchers utilized the same size class intervals when presenting their distribution results, thus allowing direct comparisons between the datasets to be made.

Table 1 presents the percentage of respiratory droplets at origin (i.e. leaving the mouth) by size class for each of the four datasets. These represent the average number of droplets (as a percentage of the total) in each size class for an individual either coughing or speaking. With the exception of Xie et al [31], who reported the percentages for each size class, the percentage values presented in Table 1 were computed directly from the quantitative data reported in the various study papers [21, 29-31]. The values shown for Loudon and Roberts [29] were adjusted using the methodology presented by Nicas et al [22], which assumed that the diameter of droplets at origin was twice that of the diameter measured remotely on sample plates. In order to match Loudon and Roberts’ results to the size classes used by the other researchers, we interpolated their results and mapped these onto the respective intervals shown in Table 1.

**Table 1.** Percentage of respiratory droplets by size class at origin derived from experimental results by Loudon and Roberts [29], Duguid [21], Chao et al. [30], and Xie et al. [31].

| Size Range (µm) | Size Class (µm) | Loudon & Roberts [29] Speaking (n) | Loudon & Roberts [29] Cough (n) | Duguid [21] Speaking (n) | Duguid [21] Cough (n) | Chao et al. [30] Speaking (n) | Chao et al. [30] Cough (n) | Xie et al. [31] Speaking (n) | Xie et al. [31] Cough (n) |
| --- | --- | --- | --- | --- | --- | --- | --- | --- | --- |
| 1–2 | 1.5 | NA | NA | 0.4 | 1.0 | NA | NA | 0.0 | 0.0 |
| 2–4 | 3.0 | 2.8 | 24.5 | 5.2 | 5.8 | 2.8 | 3.6 | 0.0 | 0.0 |
| 4–8 | 6.0 | 1.6 | 10.3 | 20.6 | 19.5 | 44.3 | 50.0 | 0.1 | 0.0 |
| 8–16 | 12.0 | 6.4 | 1.4 | 31.0 | 32.2 | 15.2 | 18.5 | 0.3 | 0.0 |
| 16–24 | 20.0 | 14.1 | 6.3 | 15.9 | 17.5 | 7.9 | 6.1 | 0.1 | 0.3 |
| 24–32 | 28.0 | 19.7 | 13.5 | 9.5 | 8.5 | 5.3 | 2.3 | 0.2 | 0.0 |
| 32–40 | 36.0 | 17.3 | 13.2 | 4.8 | 4.8 | 2.6 | 2.2 | 0.2 | 0.5 |
| 40–50 | 45.0 | 14.5 | 12.3 | 2.4 | 2.2 | 2.8 | 1.8 | 8.9 | 6.2 |
| 50–75 | 62.5 | 10.9 | 8.7 | 2.8 | 2.8 | 3.0 | 1.8 | 51.1 | 30.8 |
| 75–100 | 87.5 | 4.8 | 4.7 | 2.0 | 1.7 | 2.2 | 1.3 | 19.3 | 23.4 |
| 100–125 | 112.5 | 2.4 | 1.8 | 1.6 | 1.0 | 2.8 | 1.5 | 8.3 | 15.0 |
| 125–150 | 137.5 | 1.1 | 0.9 | 1.2 | 0.8 | 2.6 | 1.5 | 4.6 | 5.9 |
| 150–200 | 175.0 | 2.4 | 0.6 | 0.8 | 0.7 | 2.8 | 4.0 | 4.0 | 7.4 |
| 200–250 | 225.0 | 1.2 | 0.7 | 0.4 | 0.6 | 2.5 | 2.3 | 1.3 | 3.4 |
| 250–500 | 375.0 | 0.6 | 0.6 | 1.2 | 0.7 | 2.3 | 1.9 | 1.6 | 4.5 |
| 500–1000 | 750.0 | 0.1 | 0.3 | 0.4 | 0.2 | 0.8 | 1.3 | 0.1 | 2.4 |
| 1000–2000 | 1500.0 | 0.2 | 0.3 | 0.0 | 0.0 | 0.0 | 0.0 | 0.0 | 0.2 |
| Reported average number of droplets per person (n) | NA | 1765 | 465 | 252 | 4973 | 4539 | 947 | 760 | 800 |

Legend: NA – Not applicable

### 2.1 Droplet analysis

Analysis of the datasets above was undertaken using in-house algorithms written in R (R Core Team (2013). R: A language and environment for statistical computing. R Foundation for Statistical Computing, Vienna, Austria. URL http://www.R-project.org/). For each dataset the number of particles at origin in the various size classes was computed. Because initial infection with COVID-19 is thought primarily to involve cells in the nasal mucosae and the upper-respiratory tract [32, 33] as opposed to those in the lower respiratory tract, we were not only interested in droplet nuclei <10 μm diameter, but also in particles μ10 m that might become suspended in air. These when inhaled, would tend to impact on the nasal turbinates and cells in the upper-respiratory tract, and as such have the potential to transmit COVID-19. To this end, we used the droplet cut-off value of <100 μm diameter proposed by Wells [12, 14] and supported by Xei et al [13] as the criteria for identifying aerosol particles capable of transmitting the disease. Droplets <100 μm diameter at origin have been shown to rapidly reduce to about 20 to 34% of their original size due to evaporation [14, 21] with the result that they can remain suspended in the air and be transported on room convection currents [6]. Using this criteria we then computed for each dataset the percentage of droplets at origin that were <100 μm in diameter.

Because variation existed between the four datasets for both the number and distribution of the respiratory droplets produced, we decided to adopt a stochastic approach to the modelling. This involved aggregating the published experimental data and computing a theoretical distribution for the total number of respiratory droplets liberated during speaking and coughing. This was done by fitting standard distributions (i.e. lognormal, Weibull and normal) to the aggregated data using the *‘fitdistrplus’* package in R [34], with the best-fit being the one that exhibited the lowest *Akaike information criterion* (AIC). Random samples from the fitted distribution were then used in the various stochastic simulations to mimic the variance in droplet production that might be encountered in real life. For each simulation, the fraction of droplets <100 μm in diameter, *f_<100_*, exhaled during a REE was computed by randomly sampling a vector containing the 100 μm diameter cut-off percentages derived from the four study datasets shown in Table 1. Having done this, the total number of droplets <100 μm in diameter, *N_<100_*, liberated in any given simulation was computed using:

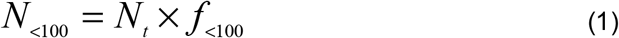

Where: *N_t_* is the number of respiratory droplets exhaled during the REE, whose value was determined by randomly sampling the best-fit distribution as described above.

In addition to calculating the numbers of particles in each size class for the four study datasets [21, 29-31], we also computed the volumetric fraction of the droplets <100 μm in diameter. This was done using the methodology described in Nicas et al [22], where the volume of the average droplet, *v_i_*, in each class or ‘bin’ was determined using:

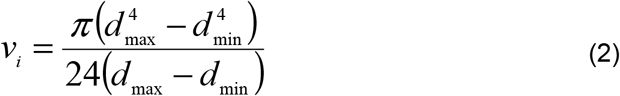

Where; *d_max_* and *d_min_* are the upper and lower diameter values (μm) in the respective size class bins.

The total volume, *V_t_*, of the droplets in each size class bin was then determined using:

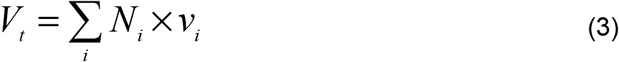

Where; *N_i_* is the number of respiratory droplets in each size class bin.

### 2.2 Model

In order to assess the airborne risk posed by a subject infected with COVID-19 occupying an enclosed space, we constructed using algorithms written in R, an *in silico* model of a 4 x 4 x 2.5 m high office space into which we placed a single infector. These dimensions were deliberately selected because they are typical of the space that might be allocated to several office workers under conditions of social distancing. In theory, if a >2 m distancing rule were observed, then it would be possible to accommodate four individuals in a space with these dimensions. However, in our study, in addition to the infector we placed just one susceptible individual in the space, who was assumed to be located >2 m away from the infector and therefore out of range of any large ballistic droplets. This individual was assumed to have a pulmonary ventilation rate (minute ventilation rate), *P_v_*, of 6 L/min (0.36 m^3^/h), which is typical for an adult performing sedentary work [35]. Analysis was performed initially using a default ventilation rate of 2 air changes per hour (AC/h) in the office space, and thereafter for a range of ventilation rates from 1-10 AC/h.

In the model we assumed that all particles <100 μm diameter liberated by a REE would quickly deduce in size due to evaporation to form small aerosol particles [13, 25] suspended in the air. Respiratory droplets >100 μm diameter were assumed to fall to the floor. We also assumed that the room air was completely mixed (which in most room spaces is a reasonable approximation [36]) and that any airborne particles produced by a REE would quickly become evenly dispersed throughout the room space. Thereafter, the concentration of the particles in the room air would fall exponentially according to equation 4, as deposition and the ventilation system removed aerosol particles from the space.

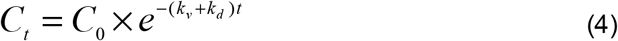

Where: *C_0_* and *C_t_* are the concentrations of respiratory particles in the air (particles/m^3^) at time zero and *t* hours respectively; *k_v_* is the room ventilation rate (air changes per hour); *k_d_* is the particle deposition rate constant (set at 5.1986 h^-1^ in accordance with experimental work by Stadnytskyi et al. [6]); and *t* is time in hours.

The total number of respiratory particles inhaled, I, by the susceptible individual was then determined using:

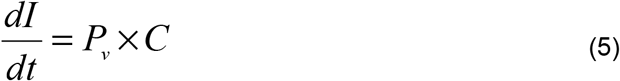

Where; *C* is the concentration of respiratory droplets in the room air (particles/m^3^).

### 2.3 Simulations

In order to quantify the risks associated with an infector located in the room space, we simulated the scenarios listed in Table 2. These scenarios were chosen because they approximated to situations that might be encountered in real life. Scenarios 1 and 2 were intended to mimic the situation where a susceptible person occupies a room space in which an infector has recently vacated. This involved an infector coughing once (Scenario 2) or speaking (Scenario 1) at time t = 0 seconds and then leaving the room space, while the susceptible individual remained present for an hour. By contrast, scenarios 3-5 were intended to simulate the situation were both the infector and the susceptible individual shared the space for an hour. Scenario 3 was intended to simulate the situation where an undiagnosed asymptomatic (or pre-symptomatic) COVID-19 infector is present in the office space. In this scenario, the asymptomatic individual performs ten speaking events (each equivalent to counting from 1 to 100) in one hour. Scenario 5 simulates the situation where a symptomatic COVID-19 infector is present in the office. In this scenario, four single coughs, equally spaced, were overlaid on the ten speaking events outlined for Scenario 3. For completeness, Scenario 4 presents the unlikely situation in which the infector coughs four times in one hour but does not speak.

**Table 2.** Details of the various scenarios simulated in the study.

| Scenario | REE type | Number of REEs per hour | Simulation duration (hours) | Description |
| --- | --- | --- | --- | --- |
| 1 | Speaking | 1 | 1 | A single 'talking event' occurring at the start of the hour. One susceptible individual present throughout. |
| 2 | Cough | 1 | 1 | A single cough occurring at the start of the hour. One susceptible individual present throughout. |
| 3 | Speaking | 10 | 1 | The same 'talking event' repeated ten times in an hour, evenly spaced. One susceptible individual present throughout. |
| 4 | Cough | 4 | 1 | The same 'cough' repeated four times in an hour, evenly spaced. One susceptible individual present throughout. |
| 5 | Speaking and coughing | 10 + 4 | 1 | Scenarios 3 and 4 combined (i.e. 10 speaking events and 4 coughs). One susceptible individual present throughout. |

Legend: REE – respiratory exhalation event

**Table 3.** Results of analysis of droplet distributions by number and volume.

|  | Loudon & Roberts [29] | Loudon & Roberts [29] | Duguid [21] | Duguid [21] | Chao et al. [30] | Chao et al. [30] | Xie et al. [31] | Xie et al. [31] |
| --- | --- | --- | --- | --- | --- | --- | --- | --- |
|  | Speaking | Coughing | Speaking | Coughing | Speaking | Coughing | Speaking | Coughing |
| Total number of particles (n) | 1765 | 465 | 252 | 4973 | 4539 | 947 | 760 | 800 |
| Total volume of particles (mL) | 0.006828 | 0.003062 | 0.000360 | 0.008352 | 0.013847 | 0.003778 | 0.000921 | 0.009552 |
| No. of particles <100 $\mu\text{m}$ (n) | 1626 | 441 | 238 | 4775 | 3909 | 829 | 609 | 490 |
| Volume of particles <100 $\mu\text{m}$ (mL) | 0.000083 | 0.000019 | 0.000004 | 0.000073 | 0.000070 | 0.000009 | 0.000108 | 0.000104 |
| No. of particles >100 $\mu\text{m}$ (n) | 139 | 24 | 14 | 198 | 630 | 118 | 151 | 310 |
| Volume of particles >100 $\mu\text{m}$ (mL) | 0.006745 | 0.003043 | 0.000356 | 0.008279 | 0.013777 | 0.003769 | 0.000813 | 0.009448 |
| Percentage of particles <100 $\mu\text{m}$ by number (%) | 92.12 | 94.84 | 94.44 | 96.02 | 86.12 | 87.6 | 80.13 | 61.25 |
| Percentage of particles <100 $\mu\text{m}$ by volume (%) | 1.19 | 0.60 | 1.07 | 0.85 | 0.49 | 0.23 | 11.66 | 1.07 |

**Table 4.**
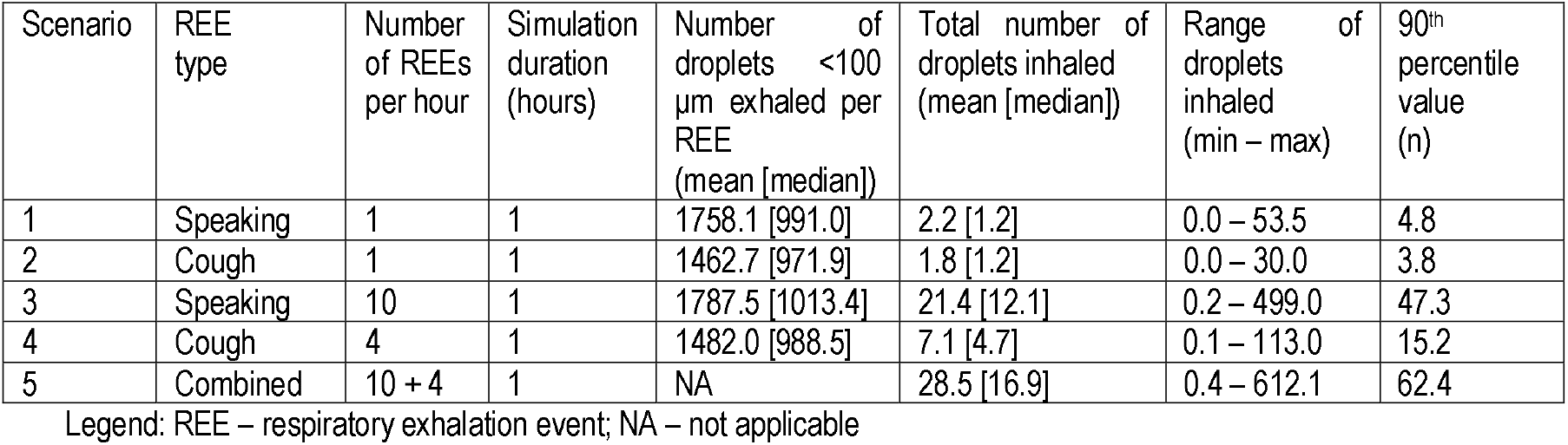
Results of office space simulations (n = 10000), assuming a default ventilation rate of 2 AC/h.

| Scenario | REE type | Number of REEs per hour | Simulation duration (hours) | Number of droplets <100 $\mu\text{m}$ exhaled per REE (mean [median]) | Total number of droplets inhaled (mean [median]) | Range of droplets inhaled (min – max) | 90<sup>th</sup> percentile value (n) |
| --- | --- | --- | --- | --- | --- | --- | --- |
| 1 | Speaking | 1 | 1 | 1758.1 [991.0] | 2.2 [1.2] | 0.0 – 53.5 | 4.8 |
| 2 | Cough | 1 | 1 | 1462.7 [971.9] | 1.8 [1.2] | 0.0 – 30.0 | 3.8 |
| 3 | Speaking | 10 | 1 | 1787.5 [1013.4] | 21.4 [12.1] | 0.2 – 499.0 | 47.3 |
| 4 | Cough | 4 | 1 | 1482.0 [988.5] | 7.1 [4.7] | 0.1 – 113.0 | 15.2 |
| 5 | Combined | 10 + 4 | 1 | NA | 28.5 [16.9] | 0.4 – 612.1 | 62.4 |

Legend: REE – respiratory exhalation event; NA – not applicable

**Table 5.**
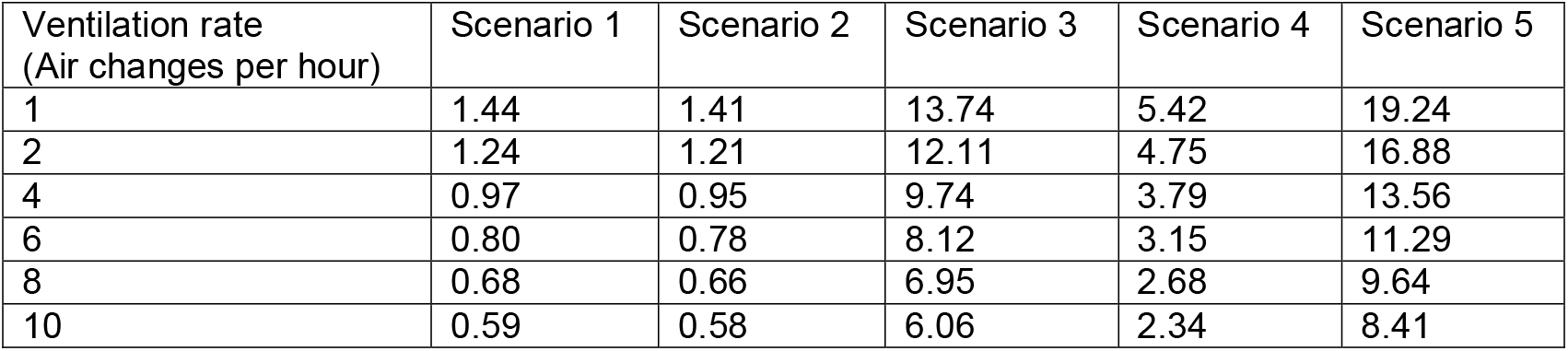
The effect of increasing the room ventilation rate on the median number of particles inhaled.

| Ventilation rate (Air changes per hour) | Scenario 1 | Scenario 2 | Scenario 3 | Scenario 4 | Scenario 5 |
| --- | --- | --- | --- | --- | --- |
| 1 | 1.44 | 1.41 | 13.74 | 5.42 | 19.24 |
| 2 | 1.24 | 1.21 | 12.11 | 4.75 | 16.88 |
| 4 | 0.97 | 0.95 | 9.74 | 3.79 | 13.56 |
| 6 | 0.80 | 0.78 | 8.12 | 3.15 | 11.29 |
| 8 | 0.68 | 0.66 | 6.95 | 2.68 | 9.64 |
| 10 | 0.59 | 0.58 | 6.06 | 2.34 | 8.41 |

For all scenarios the model simulated the total number of particles (infectious or not) that would be inhaled by a susceptible individual occupying the office space for an hour. In order to assess the variance associated with this, the simulations were repeated 10000 times so that the distribution of the likely outcomes could be assessed. For all the scenarios the number of inhaled particles was computed for a range of room ventilation rates (i.e. 1, 2, 4, 6, 8 and 10 AC/h).

## 3.0 Results

The results of the droplet distribution analysis for the four study datasets are presented in Figure 1, which shows the percentage breakdown, by size class, of the number of respiratory particles liberated at the mouth by: (i) counting from 1 to 100 (Figure 1(a)); and (ii) a single cough (Figure 1(b)). From these, and the results of the droplet analysis (Table 3), it can be seen that on average, during speaking 88.2% (range = 80.1 – 94.4%) of the droplets produced were <100 μm in diameter, whereas for coughing this figure was 84.9% (range = 61.2 – 96.0%). By comparison, most of the volume associated with the exhaled respiratory fluid resided in the larger droplets. On average, 96.4% (range = 88.3 – 99.5%) of the total fluid volume expressed during speaking was contained in droplets >100 μm in diameter, with this value rising to 99.3% (range = 98.9 – 99.8%) for coughing.

**Figure 1.**
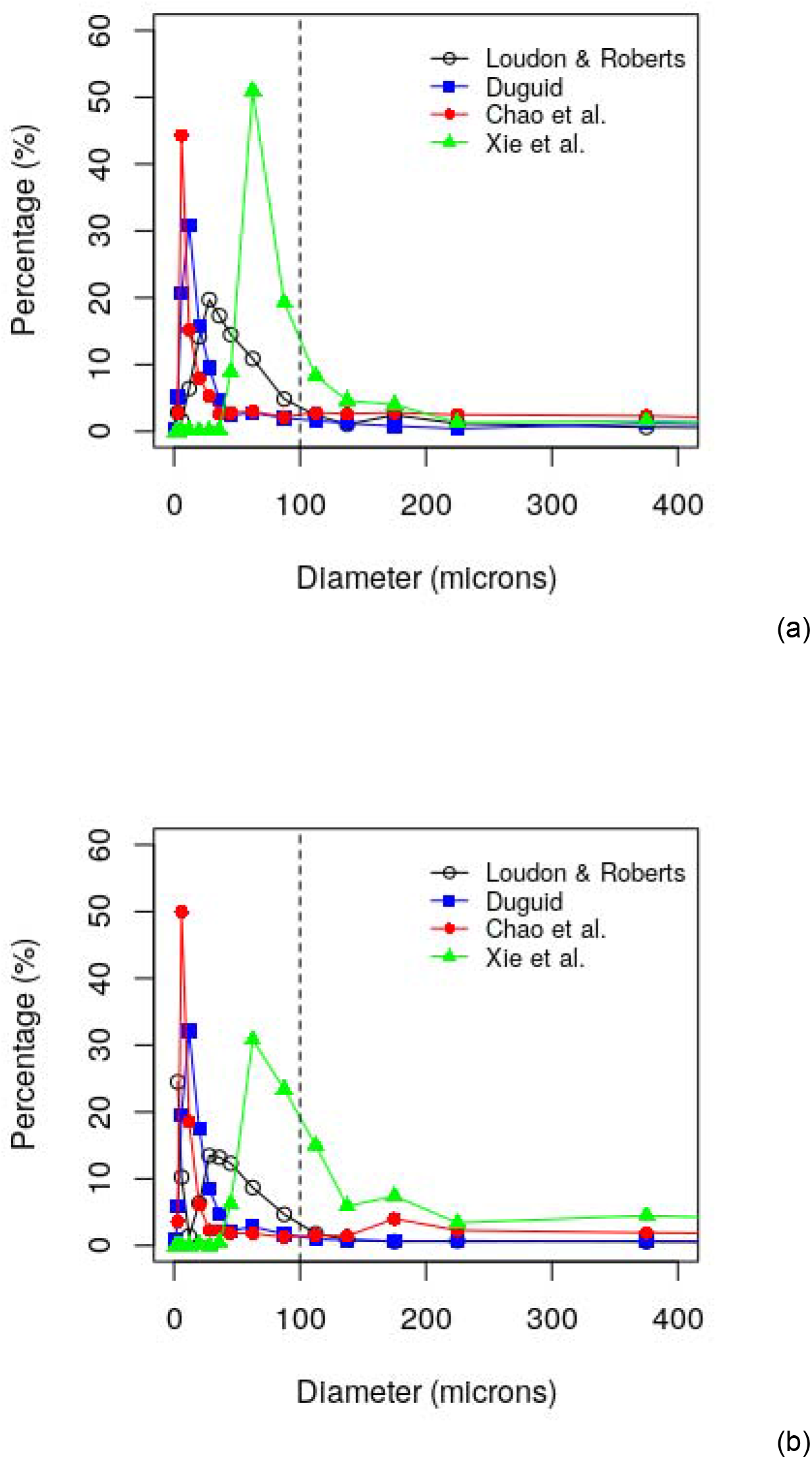
Distribution of number (expressed as a percentage of the total) of respiratory droplets at origin exhaled during: (a) counting from 1 to 100, and (b) a single cough, as reported by Loudon and Roberts [29], Duguid [21], Chao et al. [30], and Xie et al. [31]. The vertical dotted line indicates the 100 μm diameter cut-off threshold.

Analysis of the findings of Loudon and Roberts [29], Duguid [21], Chao et al. [30], and Xie et al. [31] regarding the total number of droplets liberated suggested that, although great variation exists between individuals and REEs, the distribution of the aggregated results for the total number of droplets produced broadly conformed to a lognormal function, with a geometric mean (GM) of 1113.0 and a geometric standard deviation (GSD) of 2.902 for speaking (AIC = 72.0), and GM = 1150.5 and GSD = 2.423 for coughing (AIC = 70.8). This meant that the best-fit distributions for the total number of droplets produced by the REEs were heavily positively (right) skewed, with skewness values of 0.53 and 0.73 observed for speaking and coughing respectively. So although the mean number of droplets produced during speaking and coughing were 1829 and 1796 respectively, the median values were only 1263 and 874 respectively. As such, these values appear to sit comfortably within the ranges estimated by Chao et al. [30] (i.e. 112–6720 droplets expelled during speaking and 947–2085 droplets were expelled per cough).

The results of the office space simulations using the default ventilation rate of 2 AC/h are presented in figures 2-5 and in Table 4. These reveal that spending an hour in the room space after the infector had spoken or coughed just once (scenarios 1 and 2), resulted on average (median value) in just 1.2 particles from the infector being inhaled by the susceptible individual. However, if the infector remained present in the room throughout, then this value rose to 12.1 (25-75^th^ percentile range = 5.6-25.0) particles for the asymptomatic person (Scenario 3) and 16.9 (25-75^th^ percentile range = 8.1-33.9) particles for the symptomatic person (Scenario 5).

**Figure 2.**
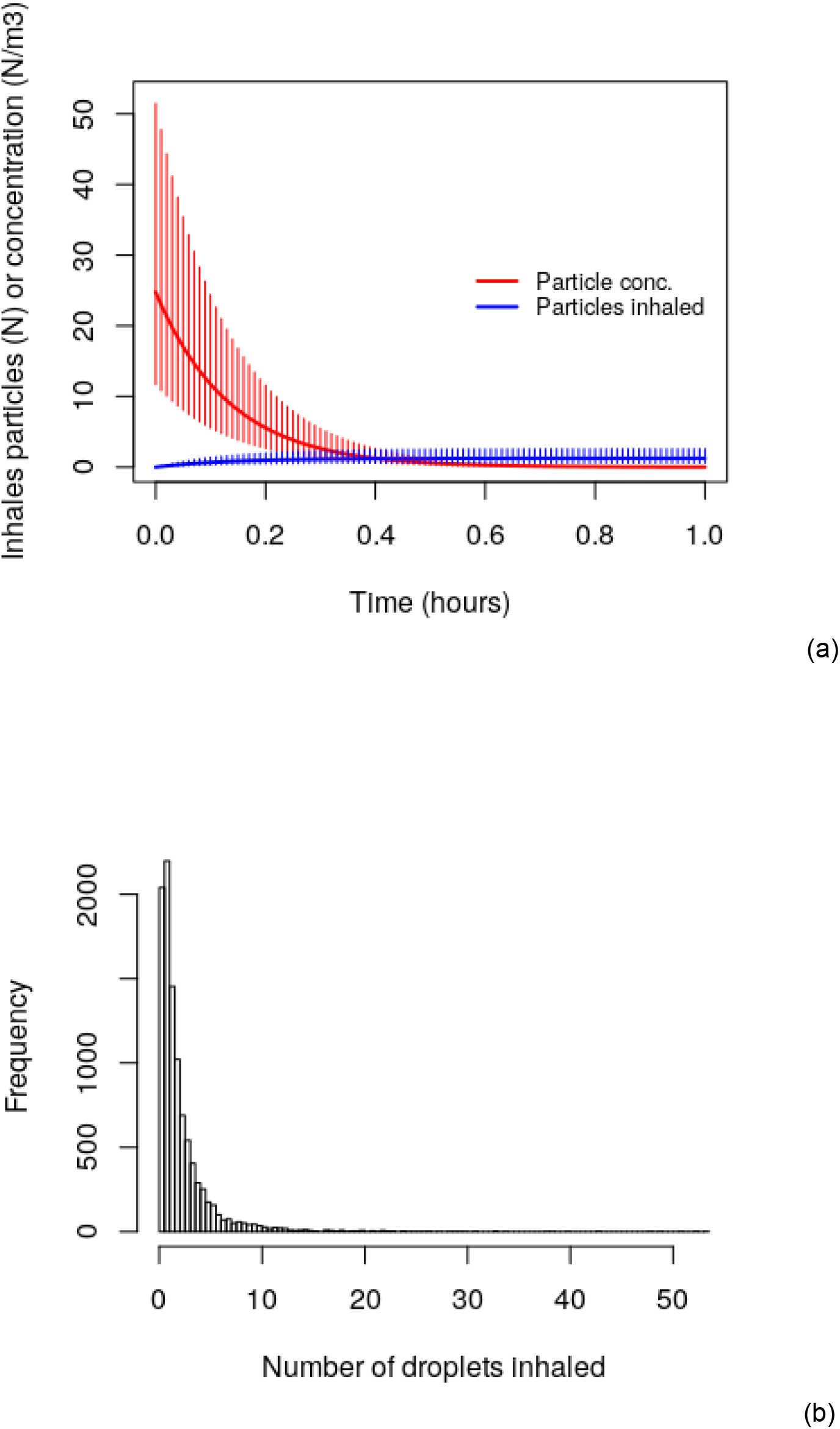
Results of the Scenario 1 ‘speaking’ simulations, showing: (a) the time plot for the median number of particles inhaled and the median particle concentration in the air; and (b) the distribution of the numbers of particles inhaled, when the ventilation rate is 2 AC/h. Results are for 10,000 simulations. Error bars represent the distribution between 25^th^ and 75^th^ percentiles.

**Figure 3.**
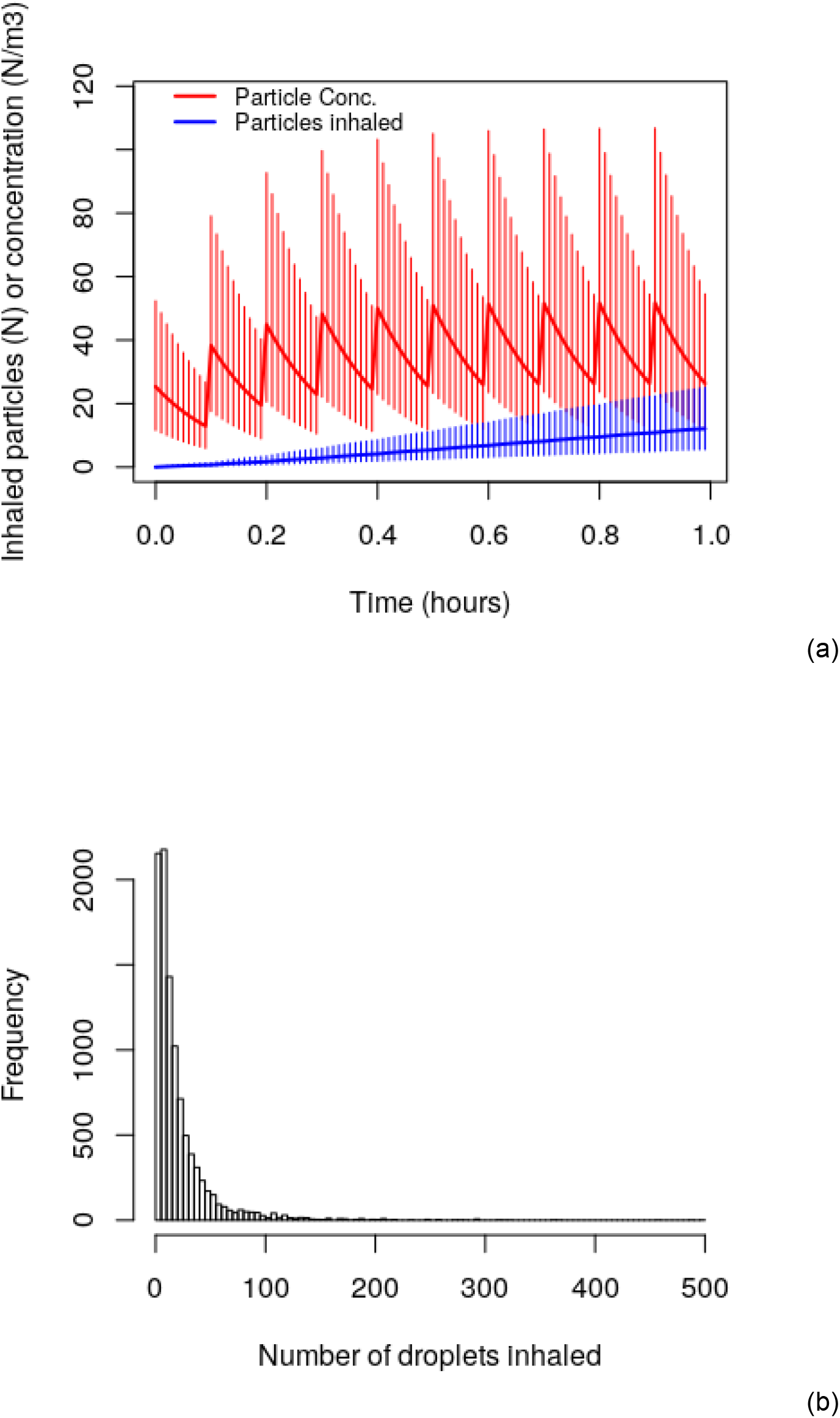
Results of the Scenario 3 ‘speaking’ simulations, showing: (a) the time plot for the median number of particles inhaled and the median particle concentration in the air; and (b) the distribution of the numbers of particles inhaled, when the ventilation rate is 2 AC/h. Results are for 10,000 simulations. Error bars represent the distribution between 25^th^ and 75^th^ percentiles.

**Figure 4.**
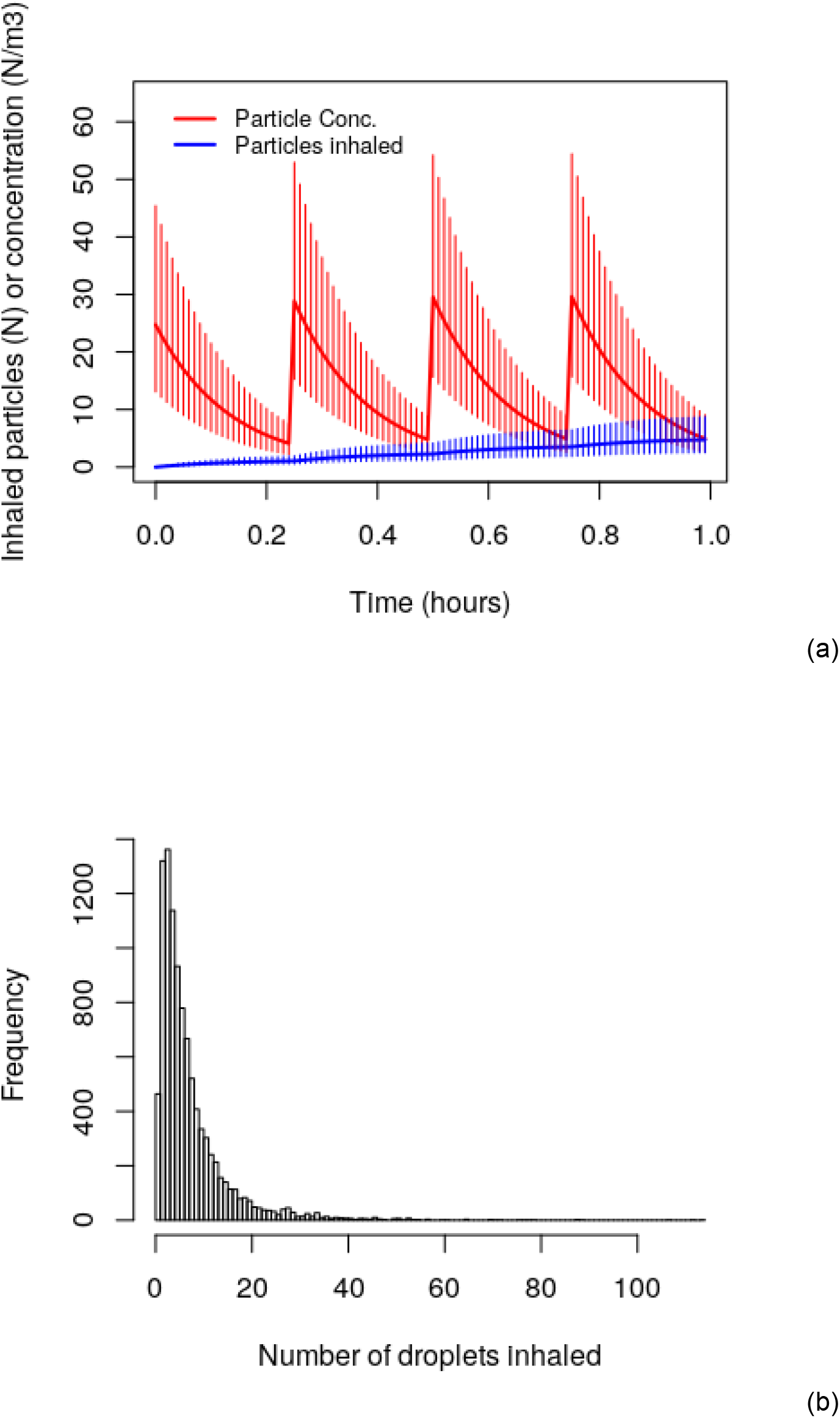
Results of the Scenario 4 ‘coughing’ simulations, showing: (a) the time plot for the median number of particles inhaled and the median particle concentration in the air; and (b) the distribution of the numbers of particles inhaled, when the ventilation rate is 2 AC/h. Results are for 10,000 simulations. Error bars represent the distribution between 25^th^ and 75^th^ percentiles.

**Figure 5.**
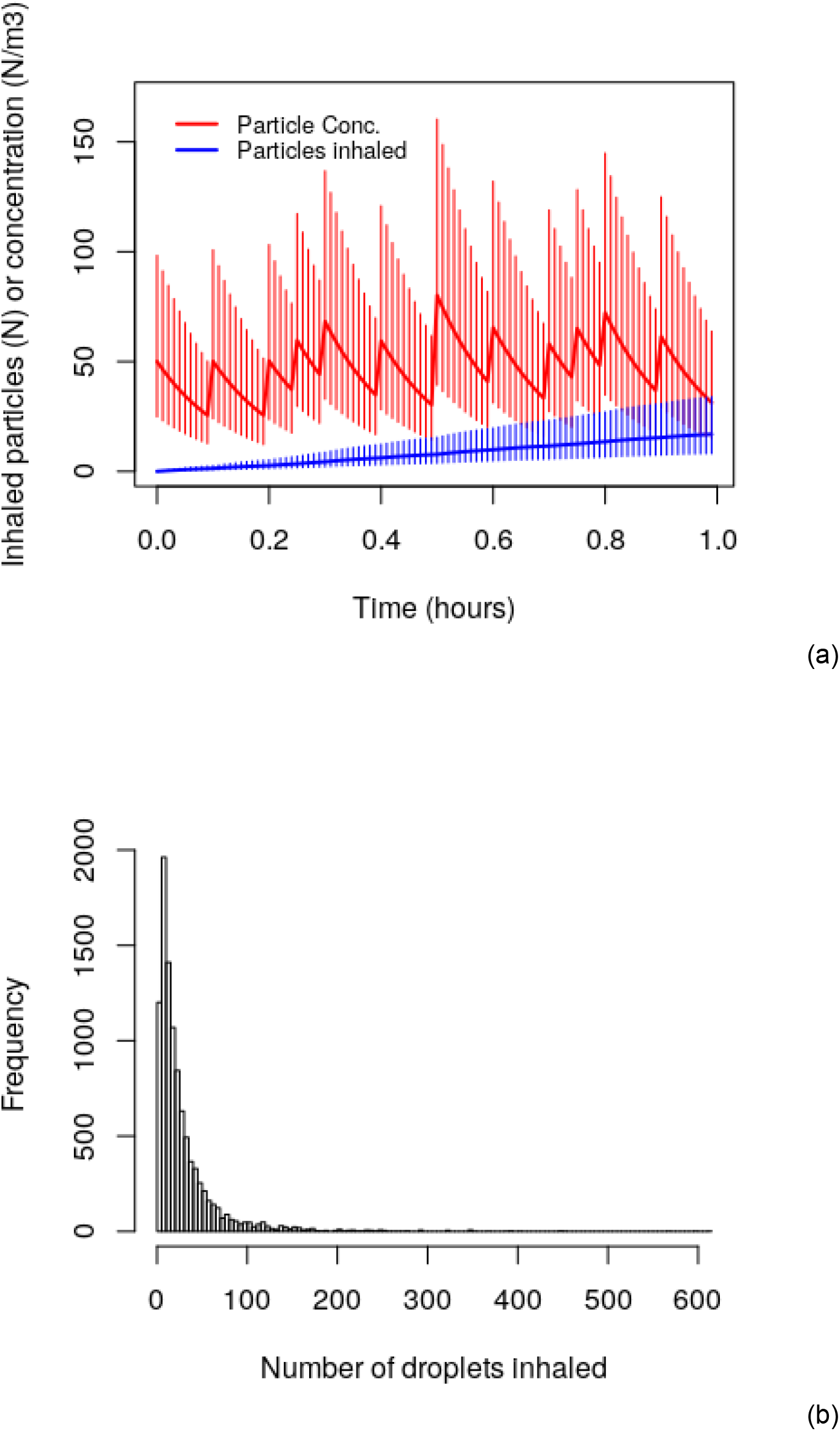
Results of the Scenario 5 ‘speaking plus coughing’ simulations, showing: (a) the time plot for the median number of particles inhaled and the median particle concentration in the air; and (b) the distribution of the numbers of particles inhaled, when the ventilation rate is 2 AC/h. Results are for 10,000 simulations. Error bars represent the distribution between 25^th^ and 75^th^ percentiles.

From figures 2(b), 3(b), 4(b) and 5(b) it can be seen that distribution of inhaled respiratory particles was highly positively skewed (skewness = 5.76; 4.90; 3.67; and 4.62 respectively). So while many of the simulations produced inhalation results that were relatively close to the median value, a few resulted in much higher inhalation values. For example, in Table 4 it can be seen that 10% of the simulations resulted in the inhalation of: >4.8 particles (Scenario 1); >3.8 particles (Scenario 2); >47.3 particles (Scenario 3); >15.2 particles (Scenario 4); and >62.4 particles (Scenario 5). As such, this suggests that a minority of individuals may inhale relatively high numbers of aerosolised respiratory droplets, particularly if they remain in the vicinity of a COVID-19 infector for long periods of time.

The impact of changing the room ventilation rate is shown in Table 5. This shows that increasing the ventilation rate resulted in an exponential reduction in the numbers of particles inhaled. However, this effect was relatively modest, with a five-fold increase in ventilation rate from 2 to 10 AC/h approximately halving the median number of particles inhaled for all scenarios.

## 4.0 Discussion

While the results presented above are constrained by the limited data available, the assumptions made, and the idealized nature of the modelling, they do however suggest that susceptible individuals will inhale appreciable numbers of respiratory particles if they spend prolonged periods of time in the vicinity of a COVID-19 infector, even if they are not directly exposed to any ballistic respiratory droplets. For example, at a ventilation rate of 2 AC/h the model indicates that spending an hour in the presence of a symptomatic COVID-19 infector (Scenario 5) is likely to result in the inhalation of 8-34 (25-75^th^ percentiles) respiratory particles that originate from the infector, which equates to approximately 280-1190 particles inhaled during a 35 hour working week. However, on 10% of occasions the number of particles inhaled exceeded 62 particles in an hour. These results are of course only an approximation and as such should be treated with caution. In reality the number of respiratory particles inhaled will depend on many factors, including: the time spent in each other’s presence; the room size; the ventilation rate; the number of times that REEs occur; and the quantity and distribution of any respiratory particles produced by each REE. Therefore, our results should be treated as a first approximation for indicative purposes only. However, notwithstanding these limitations, the results imply that if someone with COVID-19 is present in an enclosed space, others in that space may be at risk of inhaling aerosol particles containing SARS-CoV-2 virions, even when social distancing is practiced. As such, this supports the findings of other studies that suggest that the airborne transmission of COVID-19 might be taking place [3, 6].

Of course, the inhalation of respiratory droplets and droplet nuclei exhaled from COVID-19 positive individuals does not necessarily mean that any infection will be transmitted. This is because the particles inhaled may not contain any SARS-CoV-2 virions, or alternatively, the dose received may be below the threshold necessary to cause an infection in a susceptible individual. Also, the host’s immunological status might mean that they are less susceptible to acquiring a COVID-19 infection. While the minimum infectious dose necessary to cause a COVID-19 infection is not known, the ID_10_ and ID_50_ values (i.e. the doses required to cause infection in 10% and 50% of people) for SARS-CoV-1 (the causative agent in SARS) have been estimated to be 43 and 280 PFU respectively [37]. Therefore, if the infectivity of SARS-CoV-2 is similar to that of SARS-CoV-1, this implies that inhalation of just a few hundred virions might be enough to infect 50% of those exposed to COVID-19. Furthermore, it has been shown recently that the average virus RNA load in the sputum of hospitalised COVID-19 patients is 7.00 × 10^6^ copies per mL, rising to a maximum of 2.35 × 10^9^ copies per mL [38]. If we apply these values to the results presented in Table 3, we find that a COVID-19 infector would exhale on average 38423 viral copies (i.e. 0.005489 mL × 7.00 × 10^6^ copies per mL) during a speaking REE, of which 1384 viral copies would end up in aerosolised particles. Similarly, the average cough would liberate 43302 viral copies, with 298 becoming airborne. However, if we apply the maximum value for viral load stated above, then these calculated figures become approximately three orders of magnitude greater, with an estimated maximum of 464692 viral copies becoming aerosolised during a speaking REE. Given such high numbers, and the fact that the average REE results in the region of 500-2000 droplets becoming aerosolised, this suggests that many of the particles inhaled are likely to contain SARS-CoV-2 virions. As such, this appears to support Stadnytskyi et al’s [6] estimate that the probability of a 50 μm diameter droplet, prior to dehydration, containing at least one virion is approximately 37%. If the *independent action hypothesis* (IAH), which states that each virion has an equal, non-zero probability of causing an infection [39], is applicable to COVID-19 infection in humans, then in theory the inhalation of the numbers of respiratory particles per hour reported in Table 4, could be enough to seed a COVID-19 infection in a proportion of the individuals exposed. However, the extent to which the IAH is valid for humans and SARS-CoV-2 has not yet been established, although it has been shown that the IAH applies to systems where the host is highly susceptible to the disease [40]. The dose response for Middle East respiratory syndrome (MERS) in mice, which is a related disease to COVID-19, has also been shown to approximate to the IAH [41].

The results of the office space simulation highlight the crucial importance of identifying and excluding where possible COVID-19 positive individuals. Although in our simulations, the greatest number of respiratory droplets was liberated by the symptomatic COVID-19 infector (Scenario 5), it is likely that the asymptomatic infector (Scenario 3) posed a greater threat. This is because they displayed no obvious symptoms that would alert either themselves or their colleagues to any threat, despite the fact that symptomatic and asymptomatic patients have been found to display similar viral loads [42]. Consequently, it is likely that no evasive action would be taken, other than social distancing, with the result that over the course of a typical working week (35 hours), a susceptible individual sharing the office space might inhale in the region 196-875 respiratory particles originating from the infector. As such, our model suggests that undiagnosed COVID-19 positive individuals who are either asymptomatic or pre-symptomatic may play an important role in the airborne transmission of COVID-19 in buildings and other enclosed spaces, even when social distancing is observed. Indeed, given that live coronavirus is thought to be shed in high concentrations from the nasal cavity of COVID-19 positive individuals before symptoms develop [42-44], this finding is perhaps unsurprising. There is strong evidence that many infected individuals who transmit COVID-19 are either minimally symptomatic or asymptomatic [43, 45, 46], and while much of this transmission will occur through either direct contact or the action of ballistic droplets, our findings suggest that aerosolised viral particles may also be contributing to the spread of the infection. With this in mind, it is worth remembering that people behave very much like involuntary air samplers. The longer they remain within a space containing an infectious person, the more likely they are to acquire (sample) an infectious dose that might lead to disease. In the simulations shown in figures 3 and 5, we assumed that the infectious and susceptible individuals shared the same space for just one hour. In reality however, in a work place situation or, say, on a long journey in a vehicle, the two might be in the same vicinity for many hours, in which case the risk of one acquiring an infection from the other will greatly increase [47]. Similarly, if we placed multiple air samples in a room space containing an infectious person, then it would be much more likely that one or more of the samplers would return a positive result compared to the situation were a single sampler was used. In other words, if we were to place say six susceptible individuals in the office space in our simulation, then it would be much more likely that at least one might become infected within the hour. As such, this suggests that it is important to maintain low occupancy densities within enclosed spaces, as well as managing occupancy levels so that likelihood of susceptibles and unidentified infectors sharing a room space for any length of time is minimized [48].

From the results for scenarios 1 and 2 it can be seen that the room ventilation system and gravitational deposition acted together to rapidly purge the space of particles exhaled by the infector, so that within about 12-15 minutes of the infector vacating the room, the concentration of particles in the room air had fallen to relatively safe levels. From this we can deduce that the risk of inhaling infectious particles when entering an apparently empty room (from which an infector has recently exited) is usually relatively small. However, while the mechanical ventilation system was able to quickly remove any airborne respiratory droplets once the room had been vacated, the results in Table 5 reveal that it struggled to maintain safe levels when the infector was present. Furthermore, increasing the ventilation rate had only a limited effect. For example in Scenario 5, increasing the room ventilation rate from 2 to 10 AC/h only resulted in a 50% reduction in the median number of particles inhaled. As such, this suggests that relying on dilution ventilation alone to protect room occupants is a questionable strategy and that other protective measures might be required.

Unlike diseases such as TB, where infectious particles need to travel to the alveoli in order to cause an infection, COVID-19 is primarily associated with viral particles coming into contact with the angiotensin-converting enzyme 2 (ACE-2) receptors in the nasopharyngeal region and upper respiratory tract [49]. As such, inhaled particles do not need to be <10 μm in diameter in order to infect an individual, rather they can be any size. This means that larger particles, say in the region 50-10 μm, that can remain suspended in air for long enough to contaminate a mechanical ventilation system [11], can transmit the disease. Particles of this size, which often occur due to the evaporation of larger droplets [13, 25], behave as aerosol particles and can be transported on room air currents [25, 26], with the result that they can become widely dispersed, particularly if the air is turbulent. Recently, it has been shown that coughs and sneezes produce not only mucosalivary droplets but also a multiphase turbulent gas cloud that entrains the air, trapping and carrying within it clusters of droplets, both large and small, in a manner that allows the contained droplets to evade evaporation for much longer than would otherwise be the case [5]. Under these conditions, droplets of all sizes can be transported as far as 7-8 m through the air [5, 11]. This supports the findings of other researchers [13, 25, 26, 50], all of whom found that droplets >10 μm diameter could be transported >3 m. Collectively, this shows the traditional binary demarcation between droplets and droplet nuclei (<5 or 10 μm) to be an outdated concept [8-10]. Rather, every REE produces a continuum of respiratory particles, many of which can remain suspended in room air allowing them to become widely dispersed. Failure to recognise this fact, can lead to the erroneous conclusion that all ‘droplets’ (i.e. <5 μm diameter) rapidly fall to the ground, thus posing no risk to room occupants >2 m away.

One of the major problems (which is largely unrecognised) associated with using the binary cut-off of <5 μm to distinguish droplet nuclei from droplets, is that it promotes confusion and ambiguity, so that if an infection is caused by the inhalation of, say, a 20 μm diameter respiratory particle, the incident might be taken as evidence of airborne transmission by an aerosol (indoor air) scientist, whereas to a clinician, the same event would be interpreted as an example of droplet transmission. This ambiguity unfortunately all too often results in polarized views and a loss of nuance. So for example, influenza A, which shares some similar transmission characteristics to COVID-19, is classified as a disease that is spread by droplet transmission, despite the fact that numerous studies have shown that influenza RNA has been recovered from droplet nuclei <5 μm in diameter, either directly from influenza patients [15, 16, 18], or from the air in healthcare facilities [17] and other settings [51, 52]. There have also been outbreaks of influenza that have been attributed to airborne transmission. Perhaps the most notorious of these occurred in the 1970s when a commercial airplane with a 56-seat passenger compartment was delayed due to engine trouble, so that the passengers had to wait on board for 4.5 hours with no mechanical ventilation. In this incident, a single index patient, who became ill with influenza within 15 minutes after boarding the plane, managed to infect 72% of the 54 passengers on board with influenza [53]. Furthermore, Cowling et al [54], investigating influenza A outbreaks in Hong Kong and Bangkok, concluded that aerosol transmission accounted for approximately 50% of all transmission events, and Coleman and Sigler [51], sampling the air in a public elementary school, found airborne influenza A virus densities to be high enough to infect individuals within minutes. Of particular note, Coleman and Sigler also found that the viral RNA was concentrated in droplet nuclei <4 μm in diameter, rather than larger ‘droplets’, a finding mirrored by other researchers [15, 16, 18]. This is interesting because early studies on influenza in humans found that illness could be induced with substantially lower virus titers when administered as a small droplet aerosol rather than by nasal droplets, suggesting that infection is induced more efficiently when virus is deposited in the lower respiratory tract rather than the upper respiratory tract [55]. If this is the case, then it might be that with influenza, small virus laden droplet nuclei pose a greater threat than larger droplets, or at least contribute to the spread of the disease [56].

COVID-19, like influenza, is caused by an enveloped RNA virus, so it might be expected that it too will survive in the air. Indeed, it has been shown that SARS-CoV-2 can remain viable in aerosols for up to 3 hours [57]. Aerosol transmission has also been demonstrated for SARS [19, 58, 59], which is closely related to COVID-19. Consequently, there is good reason to believe that aerial dissemination of respiratory droplets <100 μm together with droplet nuclei might play an important role in the transmission of COVID-19. However, the evidence to support this is only just emerging. For example, Ong et al [60], who sampled surfaces in hospital rooms containing COVID-19 patients, found extensive fomite contamination in some patient rooms, with air exhaust outlets in particular testing positive. From this they concluded that virus-laden droplet nuclei were being transported on air currents. Air conditioning has also been implicated in an outbreak that occurred in restaurant in Guangzhou, China [27] (as discussed above), where a index patient infected five unrelated diners. In this outbreak it was shown that because the space was poorly ventilated, a contaminated recirculation envelope was created which sustained a high concentration of exhaled droplet nuclei from the index patient to diners on neighbouring tables [3]. Liu et al [61] investigating the airborne transmission of COVID-19 in hospitals in Wuhan, China, found elevated levels of SARS-CoV-2 in the air in the patient toilets and also in medical staff areas, suggesting that aerosol dissemination might be an important pathway for surface contamination. Santarpia et al [62] also observed extensive viral shedding at the University of Nebraska Medical Center, which led to extensive environmental contamination. Importantly, they found that patient room air samples were 63.2% positive by RT-PCR, with a mean concentration of 2.86 copies/L of air. Samples taken outside the patient rooms in the hallways were 66.7% positive, with a mean concentration of 2.59 copies/L of air. From this they concluded that *“viral aerosol particles are produced by individuals that have the COVID-19 disease, even in the absence of cough”*, and that *“virus-containing particles were being transported from the rooms to the hallway during sampling activities”*. Furthermore, they found that the *“personal air samplers worn by sampling personnel were all positive for SARS-CoV-2, despite the absence of cough by most patients while sampling personnel were present”*. Collectively, this suggest that transmission of COVID-19 may well be occurring by the airborne route, as well as by the droplet route, supporting the findings of our study.

## 5.0 Conclusions

In conclusion, our study suggests that the aerial dissemination of small aerosol particles containing SARS-CoV-2 virions is probably a feature of COVID-19. These particles can be suspended in air for considerable periods of time and thus can be widely dispersed. Although the analysis presented here suggest that airborne transmission of COVID-19 is likely to be occurring, particularly from asymptomatic individuals, the extent to which aerosol particles contribute to the overall burden of the disease is unclear, in part, because of ambiguity and confusion regarding the distinction between droplets and droplet nuclei. Given that unreported airborne transmission of COVID-19 may well be occurring, it is recommended that further detailed work be undertaken to inform public health policy and guidelines on occupancy levels in buildings and other enclosed spaces.

## Data Availability

The study is a computer simulation assessing the risk of COVID-19 transmission in an office space. As such, it utilizes data already published elsewhere. The sources of these data are clearly indicated and cited in the manuscript.

## Notes

### Competing Interest Statement

The authors have declared no competing interest.

### Funding Statement

The study was self funded.

### Author Declarations

Ethics approval granted by the IRB of Leeds Beckett University.

